# Improving performance of polygenic risk scores for hypertension across two ancestry groups

**DOI:** 10.1101/2025.11.05.25339527

**Authors:** Marguerite R. Irvin, Vinodh Srinivasasainagendra, Nicole D. Armstrong, Amit Patki, Ulrich Broeckel, Zhe Wang, Leslie A. Lange, Nita A Limdi, Alicia Huerta-Chagoya, Joohyun Kim, Maggie C.Y. Ng, Josep M Mercader, Hemant K. Tiwari

## Abstract

Polygenic risk score (PRS) methods are evolving, and the benefit of adding functional annotations to the variant weights has been especially promising. However, less attention has been given to how the linkage disequilibrium (LD) reference panel used affects the score performance. In the current study, we compared two Bayesian approaches, one that incorporates functional annotations (LDpred-funct) and one that does not (PRS-CS), extending these applications to the hypertension (HTN) trait across two ancestry groups (European Americans EA, and African Americans, AA). In PRS-CS we used the standard HapMap 3 LD (HM3) reference panel, as well as a modified multi-ancestry reference panel (TagIt) with better coverage of variants from multiple ancestries. Individual-level data in 1,533 EA (58% with HTN) and 8,603 AA (71% with HTN) participants from the Reasons for Geographic and Racial Differences in Stroke Study (REGARDS) was used to optimize scores across the two approaches. PRS performance metrics including R^2^ and odds ratios (OR) per standard deviation (SD) were then used to assess PRS performance in 1,270 EA (55% with HTN) and 1,896 AA (69% with HTN) participants from the Hypertension Genetic Epidemiology Network Study (HyperGEN). Among EAs in HyperGEN we observed an R^2^ of 6.0% for LD-Pred-funct and R^2^ of 7.3% for PRS-CS-TagIt versus R^2^ of 1.4% for PRS-CS-HM3. The magnitude of the OR per SD for HTN was also higher for PRS-CS-TagIt OR=2.17 (95% CI 1.65-2.85, p=3.0*10^−8^) and LD-Pred-funct OR=2.14 (95% CI 1.61-2.85, p=1.46*10^−7^) versus PRS-CS-HM3 (OR=1.40; 95% CI 10.8-1.82). Among AAs in HyperGEN, the improvements were more modest, where we observed R^2^ of 1.9% for LD-Pred-funct and R^2^ of 2.9% for PRS-CS-TagIt versus 0.7% for PRS-CS-HM3. We found that both annotations and the updated LD panel improved the scores in both ancestry groups, but did not make the scores more equitable across the groups.

**Author Summary:** Genomics can aid in risk prediction and prevention. Polygenic risk score (PRS) development and application is becoming more popular because there’s a lot of genetic data available for training PRS, and they have the potential to be useful in healthcare. However, PRS are not yet widely used in clinics, and the methods are still developing. Early PRS methods only used certain genetic markers, but newer ones use more data and better models to try to predict disease risk more accurately. Still, these tools often don’t work as well for people from underrepresented populations, which could increase health inequalities. Researchers are trying to fix this by using more up-to-date data and adding extra information about how genes function. In our study of HTN, we investigated newer approaches which made PRS more accurate for both for African American individuals and European American individuals—but they didn’t fully close the performance gap between the groups.

## Introduction

Genome-wide association studies (GWAS) have uncovered a great deal about the polygenic architecture of common complex disorders. Although GWAS results have been largely challenging to translate into clinical settings, applications involving polygenic risk scores (PRS) show promise for clinical utility.(1) Methods for PRS are evolving, and standard pipelines for derivation and implementation have been proposed but are not yet established.(2) Other challenges include the varying performance of PRS across populations with diminished predictive capacity in minority populations.(3) This is most likely due to linkage disequilibrium (LD) patterns that vary across populations, and the overrepresentation of European Ancestry individuals (EAs) in genetic databases leveraged to create PRS. Strategies for improving performance are emerging but may still leave some ancestry groups behind. (4)

Currently, there is no standard method for polygenic prediction. Common approaches include pruning and thresholding (P+T) which sums the effect size of variants filtered on LD and evidence of association.(5) Alternatively, Bayesian polygenic prediction methods aim to capture variants genome-wide and, thus, do not attempt to identify a minimal subset of variants, but shrink their effects based on their LD.(6-9) These methods use prior distributions paired with advanced modeling techniques, GWAS summary statistics, and an external LD reference panel to estimate the posterior mean effect sizes. For instance, the PRS-CS method uses continuous shrinkage priors to enable SNP-specific adaptive shrinkage while allowing for diverse underlying genetic architectures across SNPs.(6) Another widely used method, LDPred, applies a point-normal prior for the SNP effect sizes after assuming there is an optimized proportion of causal variants for the trait of interest. (8, 10) Functional annotations can help prioritize more causal variants and LDPred has been extended to LDpred-funct by adding functional annotation information to the priors to estimate the mean causal effect sizes.(11)

While many comparisons have been made between PRS development approaches, the benefit of adding functional annotations to available methods is becoming increasingly evident. As functional annotations may be more likely to differentiate causal SNPs from non-causal SNPs in high LD, they may help harmonize translation performance across ancestral groups. (12) Less attention has been given to the impact the chosen LD reference panel has on the variant’s selection process. Expanding the variants available in the leveraged multi-ancestry reference panel may improve PRS performance by better capturing linkage disequilibrium patterns across diverse populations. In the current study, we compared LDpred-funct performance to PRS-CS, which does not include functional annotations, extending these applications to hypertension (HTN) across two ancestry groups (European Ancestry, EA and African Ancestry, AA). In the PRS-CS program, we utilized the standard HapMap 3 (HM3) LD reference panel, as well as a modified multi-ancestry reference panel (TagIt) with a better coverage of variants from multi-ancestry populations. (13) Our study assessed these methods using individual-level data in 1,598 EA and 8,603 AA participants with GWAS from the Reasons for Geographic and Racial Differences in Stroke Study (REGARDS) and 1,270 EA and 1,896 AA participants with GWAS from the Hypertension Genetic Epidemiology Network Study (HyperGEN).(14, 15) For each approach, REGARDS served as the optimization cohort and HyperGEN as the validation cohort.

## Results

Demographic data for the testing and validation cohorts is provided in **Table 1**. REGARDS participants on average were older than HyperGEN participants. Both cohorts had a higher percentage of females and larger case versus control counts due to the older (REGARDS) and hypertensive population (HyperGEN) sampling. The count of variants per score are presented in **Table 2** with fewer variants included for the PRS-CS method (∼800K-2M variants) and close to three times more variants represented in scores generated by LD-Pred-funct (≥3.5M variants).

**Table 1:** Cohort Characteristics.

| **Table 1: Cohort Characteristics** |  |  |  |  |
| --- | --- | --- | --- | --- |
| **Cohort** | **Age (±SD)** | **Sex (% female)** | **N HTN Case** | **N HTN Control** |
| **Optimization** |  |  |  |  |
| REGARDS AAs | 63.59 (9.20) | 60.98% | 6146 | 2457 |
| REGARDS EAs | 68.34 (10.38) | 42.30% | 896 | 637 |
| **Validation** |  |  |  |  |
| HyperGEN AAs | 47.01 (12.81) | 63.52% | 1315 | 581 |
| HyperGEN EAs | 49.83 (13.94) | 50.47% | 697 | 573 |
| AA-African American, EA-European American, HTN-Hypertension |  |  |  |  |

**Table 2:**
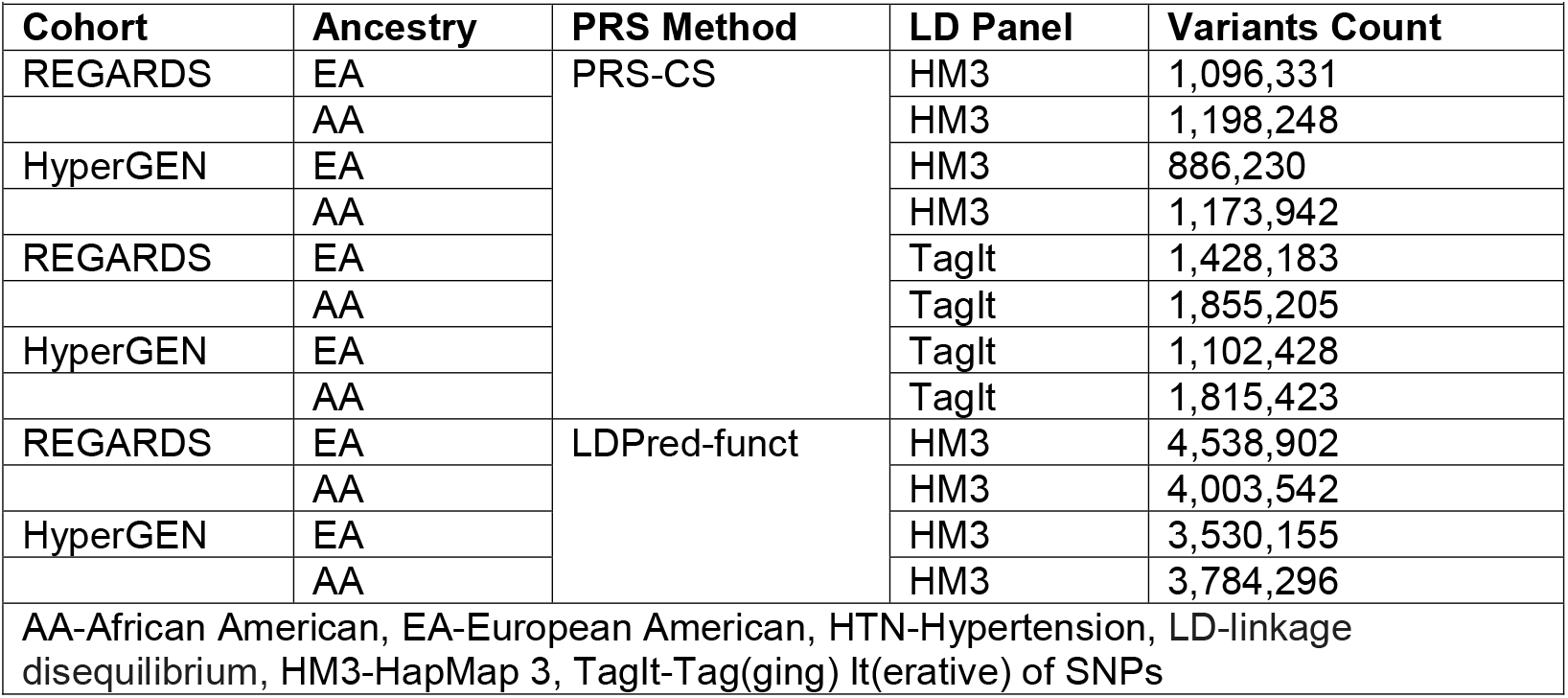
Variants Count by Method and LD Panel.

The results represented as OR per SD of the PRS optimized in REGARDS and tested in HyperGEN are presented in **Table 3**. Based on R^2^ the Phi of 1×10^−04^ was tested in PRS-CS in HyperGEN. Effect sizes and R^2^ were higher in EAs compared to AAs in both REGARDS and HyperGEN across each method. For instance, the OR for LD-Pred-funct in HyperGEN EAs was 2.14 (95% CI: 1.61-2.85) versus 1.37 (95% CI: 1.19-1.58) for AAs. In both ancestry groups, scores created by adding functional priors (LDPred-funct) and expanding the reference panel (PRS-CS-TagIt) had improved R^2^ compared to PRS-CS-HM3. We observed R^2^ of 6.0% for LD-Pred-funct and R^2^ of 7.3% for PRS-CS-TagIt in EAs from HyperGEN versus R^2^ of 1.4% for PRS-CS-HM3 (**Table 3**). Among AAs in HyperGEN, the improvements were more modest, where we observed R^2^ of 1.9% for LD-Pred-funct and R^2^ of 2.9% for PRS-CS-TagIt versus 0.7% for PRS-CS-HM3. See **Figure 2** for comparisons of R^2^ by model by cohort. See **Supplementary Table 1** for the full results for each model for each cohort.

**Table 3:** HTN Results per 1 standard Deviation of the PRS.

| **Table 3: HTN Results per 1 standard Deviation of the PRS** |  |  |  |  |  |  |
| --- | --- | --- | --- | --- | --- | --- |
| **Cohort** | **PRS Method** | **Parameter** | **OR (95% CI)** | **P** | **$R^2$** | **AUC PRS** |
| REGARDS<br>EAs | PRS-CS-HM3 | $1 \times 10^{-04} *$ | 1.37 (1.23-1.53) | 1.02E-08 | 2.71E-02 | 0.58 |
| | PRS-CS-TagIt | $1 \times 10^{-04} *$ | 1.73 (1.54-1.94) | 4.48E-21 | 7.79E-02 | 0.64 |
|  | LDPred-funct | - | 1.51 (1.35-1.68) | 3.74E-13 | 4.46E-02 | 0.61 |
| HyperGEN<br>EAs | PRS-CS-HM3 | $1 \times 10^{-04} *$ | 1.40 (1.07-1.82) | 1.19E-02 | 1.37E-02 | 0.56 |
| | PRS-CS-TagIt | $1 \times 10^{-04} *$ | 2.17 (1.65-2.85) | 3.00E-08 | 7.25E-02 | 0.61 |
|  | LDPred-funct | - | 2.14 (1.61-2.85) | 1.46E-07 | 6.03E-02 | 0.60 |
| REGARDS<br>AAs | PRS-CS-HM3 | $1 \times 10^{-04} *$ | 1.25 (1.19-1.32) | 1.39E-16 | 1.76E-02 | 0.56 |
| | PRS-CS-TagIt | $1 \times 10^{-04} *$ | 1.28 (1.21-1.35) | 5.36E-19 | 2.06E-02 | 0.57 |
|  | LDPred-funct | - | 1.32 (1.25-1.39) | 5.20E-24 | 2.66E-02 | 0.57 |
| HyperGEN<br>AAs | PRS-CS-HM3 | $1 \times 10^{-04} *$ | 1.20 (1.04-1.39) | 1.15E-02 | 6.70E-03 | 0.53 |
| | PRS-CS-TagIt | $1 \times 10^{-04} *$ | 1.47 (1.27-1.69) | 1.14E-07 | 2.94E-02 | 0.55 |
|  | LDPred-funct | - | 1.37 (1.19-1.58) | 1.76E-05 | 1.93E-02 | 0.52 |
| *Best model Phi REGARDS, AA-African American, EA-European American, HM3-HapMap 3 LD Panel, TagIt-Tag(ging) It(erative) of SNPs LD Panel, AUC-Area Under the Curve |  |  |  |  |  |  |

PRS threshold results (e.g. top 2% of the PRS distribution versus bottom 98%), are presented in **Supplementary Tables 1a** and **1b** in REGARDS EAs and AAs and **Supplementary Tables 2a** and **2b** for HyperGEN EAs and AAs, respectively. The PRS-CS-TagIt panel and LD-Pred-funct models had stronger associations with HTN at the four tested thresholds than the PRS-CS-HM3 model. For example, being in the top 5% of the PRS distribution for the score generated in LD-Pred-funct was associated with OR=8.10 (95% CI:2.48-26.51) and p=5.4×10^−04^ in HyperGEN EAs. The comparable model for PRS-CS-TagIt (phi 1×10^−04^), for the 5% threshold had OR=7.98 (95% CI: 2.27-28.00) and p=1.2×10^−03^. The AUC for the PRS-only model was 0.52 for both approaches (LD-Pred-funct and PRS-CS-TagIt) for the 5% threshold. The 5% threshold PRS-CS-HM3 model for EAs in HyperGEN was not significant (p=1.4×10^−01^). Generally, across the models in HyperGEN EAs sensitivity was low (<13.5%) and specificity was high (>0.86). The adjusted PPVs ranged 0.32-0.77 and the adjusted NPVs ranged 0.43-0.47. For HyperGEN AAs for LD-Pred-funct the 5% threshold model had an OR of 2.50 (95% CI: 1.32-4.70) with p=4.6*10^−03^. The comparable model for PRS-CS-TagIt (phi 1×10^−04^) for the 5% threshold had OR=3.50 (95% CI: 1.72-7.09) and p=5.3×10^−04^. Additionally, the sensitivities and specificities reported were consistently low and high, respectively for HyperGEN AAs (sensitivity <0.11 and specificity >0.88). ROC curves for REGARDS and HyperGEN are presented in Supplementary Figure 1.

## Discussion

PRS is increasingly popular given accessibility of large-scale GWAS summary data resources as well as the potential for broad clinical applications. Thus far, clinical translation remains limited, but methods are rapidly evolving. The first PRS approaches such as P+T subset GWAS SNPs based on different thresholds of significance and LD, while later Bayesian methods included a wider set of SNPs with more sophisticated modeling to improve the capture of trait heritability. (5, 16, 17). Current methods can be especially limiting for underrepresented groups and revised PRS development strategies are needed to ensure that new health disparities are not created in implementing PRS.(17) Adding functional annotations may help prioritize variants that are more consistent across ancestry groups.(18) Further, the standard approaches have also relied on older LD reference panels that may not capture an accurate representation of LD for diverse groups.(6, 11) In the current study we evaluated both functional annotations and an updated reference panel for improved performance of PRS for HTN, a common, heritable chronic disease in both EA and AAs. We found that both annotations and the updated panel improved the scores in both race groups, but did not make the scores more equitable across the groups.

The accuracy of PRS depends on several factors including the SNP selection and the truthfulness of their estimated effect sizes. Given evidence suggesting that common causal variants are likely to be shared across populations, it’s been hypothesized that functional annotations can be used to discriminate probable causal SNPs from other SNPs in high LD with them for better genomic prediction. (11, 19-23) However, prioritizing these variants is challenging because numerous common causal variants in LD, often with modest effect sizes, underly complex traits. (24, 25) In our study the scores created by LDPred-funct had many more variants (∼3X more) than for PRS-CS which could have diluted the effects for more important SNPs. For multi-ancestry and cross-ancestry prediction, the accuracy is additionally influenced by how well the LD in the GWAS summary statistics population and the LD panel match the optimization population. A mismatch can be an important contributor to the loss in PRS prediction accuracy in cross and multi-ancestry settings. (26) Therefore, in addition to functional annotations we also considered how updating the reference panel to a more diverse set of variants/samples could improve PRS for HTN. To date, a popular Bayesian method has used HM3 variants in 1000G LD data to construct PRS.(6) This was mostly aimed at managing computational resources while capturing overrepresented European ancestry genetic variation from large-scale GWAS summary data. However, HM3 variants are known to miss signals specific to other ancestry groups. We tested whether an expanded reference panel that includes a more diverse set of 1000G variants may overcome some of those issues by leveraging the PRISM Consortium TagIt panel in PRS-CS that captures >1 million more SNPs than the PRS-CS HM3 default panel.(13, 27) Our results show expanding the reference panel and functional annotations both improved performance to a comparable degree in EAs. Data on changing out the LD reference panel in PRS derivation is mixed given expansion.(11-13) One study showed increasing the sample size of the representative LD reference dataset linearly improved R^2^.(12) In the context of T2D in the paper by Huerta-Chagoya et. al. PRS created with the TagIt panel always outperformed scores created with the HM3 panel across 5 major ancestry groups (African/African American (AFR), Admixed American (AMR), East Asian (EAS), European (EUR), and South Asian (SAS)). Given the HM3 and TagIt panels should work well in EAs, we considered the improved performance of the TagIt panel over the standard HM3 panel in PRS-CS could be due to better representation of HTN variants in the TagIt panel. However, given the complexity and large number of SNPs in the PRS fully characterizing how differences in HTN SNP representation drive performance was beyond the scope of our project. Unfortunately, neither an expanded reference panel or functional annotations improved PRS performance much in AAs, highlighting an important trend that PRS generally are less accurate in minority populations. This is likely mostly driven by underrepresentation in the summary statistic data and the benefits of a more diverse LD reference panel may not be fully observed until training data becomes more representative.

Multi-ancestry HTN PRS have largely been trained on continuous-trait BP summary statistics for SBP and DBP, rather than solely on summary statistics for dichotomous HTN, as we did in this study.(28) In one recent study, Kurniansyah *et al*. trained a HTN PRS using GWAS summary statistics from over 50,000 diverse samples and a P+ T approach using GWAS data for SBP, DBP, and HTN, respectively.(29) A PRS was optimized for each trait and the trait-specific PRSs were joined via an unweighted sum called “PRSsum” estimating the HTN-PRS. In the validation phase of that study using over 30,000 participants from BioMe, the HTN-PRS had an OR=1.32 per SD (95% CI: 1.2-1.4) for prevalent HTN in 8723 AAs and OR=1.47 per SD (95% CI: 1.4-1.5) in 22463 EAs. In another study, that same PRS had OR=1.27 per SD (95% CI 1.2-1.3) for prevalent HTN in a combined race sample of 98,590 from the All of Us network (∼20% AA). (28) Overall, there has been a lot of research focused on optimizing PRS for HTN in diverse groups. In a multi-ancestry investigation of multiple BP PRS created by “clumping-and-thresholding” (PRSice2), LDPred2 as well as PRS-CSx (a multi-ancestry modification of PRS-CS), the PRS-CSx approach performed best across all race/ethnic backgrounds. They showed that R^2^ values for BP in AA participants were comparable to estimates reported in our study ∼3-3.5%.(28) Generally, the scores we developed in this study only improved marginally on previous publications. The body of evidence shows multi-trait training data, multi-ancestry representation in the summary data, modeling functional annotations, and improvements to the reference LD panel all increase HTN PRS performance in EAs and AAs.(28, 29) As summary statistic datasets grow and diversify, annotations improve, and computing resources expand PRS, the gap in PRS performance between AAs and EAs for HTN should narrow.

This study developed PRS for HTN in well-characterized cohorts with EA and AA adults with relevant genetic data from the REGARDS study. The scores developed were then tested in a familial study of HTN (HyperGEN) with comparable phenotypic and genetic data resources. We leveraged a novel LD reference panel developed by the PRISM Consortium, an NIH-funded program that investigates the transportability of PRS. Limitations included that the multi-ancestry summary statistics were predominantly derived from the UKB EA population, although that dataset does have samples from multiple ancestry groups. In our testing cohort, HyperGEN, some of the thresholded models had smaller participant counts (e.g. only 12 controls were in the top 5% for EAs) which likely inflated some of the OR estimates and made confidence intervals around those estimates wide. Finally, given we only studied the EA and AA populations, our work may not be generalizable to other ancestry groups.

In summary, we constructed multiple PRSs for HTN using Bayesian approaches incorporating millions of SNPs for risk prediction. We showed that updating the reference panel in PRS-CS and adding functional priors in LDPred-funct can improve HTN PRS performance. While validation of the scores showed comparability and/or marginal improvement over published data, the results still illustrate the difficulty of constructing PRS that can be clinically informative. Expanding non-European genomic resources while leveraging the growing publicly available annotation data, especially at the tissue and cell level is needed to continue to move the field forward. Further, studies linking PRS to incident HTN will be necessary to evaluate if scores for this common condition can aid in prevention and treatment.

## Methods

### Study populations

REGARDS is a national cohort established for the study of incident stroke and associated risk factors. Between 2003 and 2007 over 30,239 AA and EA adults aged 45 years or older were enrolled from all 48 contiguous US states and the District of Columbia.(30) At baseline, participants completed a computer-assisted telephone interview followed by an in-home visit that assessed blood pressure and other clinical measurements. REGARDS is ongoing and since its inception, participants have been contacted at six-month intervals to obtain information regarding incident stroke and secondary cardiovascular outcomes. HyperGEN was a cross-sectional, population-based study designed to identify genetic risk factors for HTN and target end-organ damage due to HTN.(31) The cohort, composed of self-reported AA and EA sibships, was recruited from five field centers across the US (Forsyth County, NC; Minneapolis, MN; Framingham, MA; Salt Lake City, UT; and Birmingham, AL). Recruitment included sibling probands who were diagnosed with HTN before age 60, their unmedicated adult offspring, and age-matched controls. In a later recruitment phase other siblings of the original sibling pair as well as their children were included for a total sample size of N=4584.

Informed consent was obtained from all subjects involved in the HyperGEN and REGARDS studies. The REGARDS study was conducted according to the guidelines of the Declaration of Helsinki and approved by the Institutional Review Board of the University of Alabama at Birmingham (UAB IRB protocol number: IRB-020925004). The HyperGEN study was approved by the same IRB (UAB IRB protocol number: IRB-160331004). The PRS research leveraging these datasets was approved by UAB IRB protocol number: IRB-300006721.

### Genotyping

A subset of 10,788 AA and EA REGARDS participants underwent genotyping using the Illumina Infinium AMR/AFR (MEGA) BeadChip array. Quality control procedures have been previously described, but briefly, internal duplicates and HapMap controls were excluded, as well as samples whose genetic sex did not match the self-reported sex.(14) Variants were excluded if they were located on sex chromosomes, had ambiguous strands, were not bi-allelic, were in violation of Hardy Weinberg (p<1.00E-12 for AAs and p<1.00E-5 for EAs), and/or had a missing rate >10%. Imputation in each ancestry separately was performed using the Trans-Omics for Precision Medicine (TOPMed) release 2 (r2, Freeze 8) reference panel. (32) Imputed variants were kept if r^2^ >0.3 and genotypic probability >0.9. After QC a total of 6,146 AA HTN cases and 2,457 AA controls contributed data to the analysis along with 896 and 637 EA HTN cases and controls from REGARDS.

Genotyping of AA HyperGEN participants was performed through whole genome sequencing (N=1,896).(33) In order to harmonize our imputation efforts with the array-based panels of REGARDS, we subset common bi-allelic SNPs with MAF >1% overlapping with the GWAS MEGA arrays used in REGARDS. This yielded a total of 2,204,415 SNPs that were used as fence post variants for imputing the HyperGEN AA cohort using version r2 of the TOPMed reference panel. Imputed variants were inspected for their imputation quality scores and retained if r^2^ >0.3 along with genotypic probability >0.9. HyperGEN EAs (N=1358) were genotyped with the Affymetrix Genome-Wide Human SNP Array 6.0 (1.8 million variants).

Similarly to REGARDS, internal duplicates and HapMap controls were excluded, as well as samples whose genetic sex did not match the self-reported sex. Variants were excluded if they were located on sex chromosomes, had ambiguous strands, were not bi-allelic, were in violation of Hardy Weinberg, and/or had a missing rate >10%. Imputation was performed using the TOPMed r2 (Freeze 8) reference panel.(32) Imputed variants were retained if r^2^ >0.3) and calls had genotypic probability >0.9. After QC a total of 1315 and 581 AA HTN cases and controls contributed data to the analysis, along with 697 and 573 EA HTN cases and controls from HyperGEN.

### Phenotyping

The measurement of blood pressure (BP) in the REGARDS study has been described.(34) Systolic blood pressure (SBP) and diastolic blood pressure (DBP) were defined as the average of two measurements taken during the in-home visit by a trained technician using a standardized protocol and regularly tested aneroid sphygmomanometer after the participant was seated for five minutes. HTN was defined as SBP ≥140 and/or DBP ≥90 mmHg and/or treatment with antihypertensive medication. The use of anti-hypertensive medication was self-reported. In HyperGEN, BP measurements were made during the study visit, three times on each arm. The participants rested for five minutes before measurement and 30 seconds between measurements.(31) An average SBP and DBP were calculated based on the second and third measurements in the second arm. Blood pressure measurements were obtained using automated Dinamap devices (model 1846 SX/P, Critikon, Tampa, FL) to promote comparable measurements among HyperGEN field centers. HTN was defined as average SBP ≥ 140 mmHg or average DBP ≥ 90 mmHg, or using at least one class of antihypertensive medication.

### PRS construction and optimization

The Bayesian methods applied require GWAS summary statistics, participant individual level data for optimization and an external LD reference panel for PRS construction.(6, 11) We leveraged Pan UK Biobank Multi-Ethnic GWAS summary statistics for HTN representing multi-ancestry data from 420,473 European (EUR) individuals, 6,624 African (AFR) individuals, 8,849 South Asian (SAS) individuals, 2,707 East Asian (EAS) individuals, 998 Admixed American (AMR), and 1,595 middle eastern ancestry individuals (MID).(35) For each PRS software package, the REGARDS cohort served as the optimization cohort and HyperGEN as the validation cohort.

We first applied PRS-CS using the HapMap3 (HM3) SNPs (default PRS-CS-HM3 LD panel) [20] and a newly developed TagIt LD panel from the D-PRISM Consortium (developed using the Tag(ging) It(erative) of SNPs in multiple populations (TagIt) program), designed to enhance variants available for cross-ancestry prediction through leveraging variants and samples from the 1000 Genomes project (PRS-CS-TagIt).(13, 27, 36, 37) The HM3 LD panel in PRS-CS had 1,225,319 variants, whereas the expanded TagIt panel had 2,374,614 variants with 729,110 overlapping between the two panels (6). To optimize the global scaling parameter ϕ, we applied a grid search across four ϕ values (1.0, 1×10^−02^, 1×10^−04^, 1×10^−06^) in the optimization sample. In addition, we applied LDpred-funct, which is distinct from PRS-CS since it leverages trait-specific functional priors for coding, conserved, regulatory, and LD-related annotations.(11) LDpred-funct then uses both the functional priors and information on the LD between variants from a 1000G phase 3 reference panel (including 1,217,312 variants) to estimate the posterior mean causal effect sizes. To improve computational efficiency, we ran the LDpred-funct program chromosome by chromosome. This allowed us to calculate weights at the chromosomal level and then combine them for validation. As an input parameter, SNP-heritability was calculated using LD score regression (LDSC). (38) See **Figure 1** for an outline of our pipelines for each PRS development approach.

**Figure 1.**
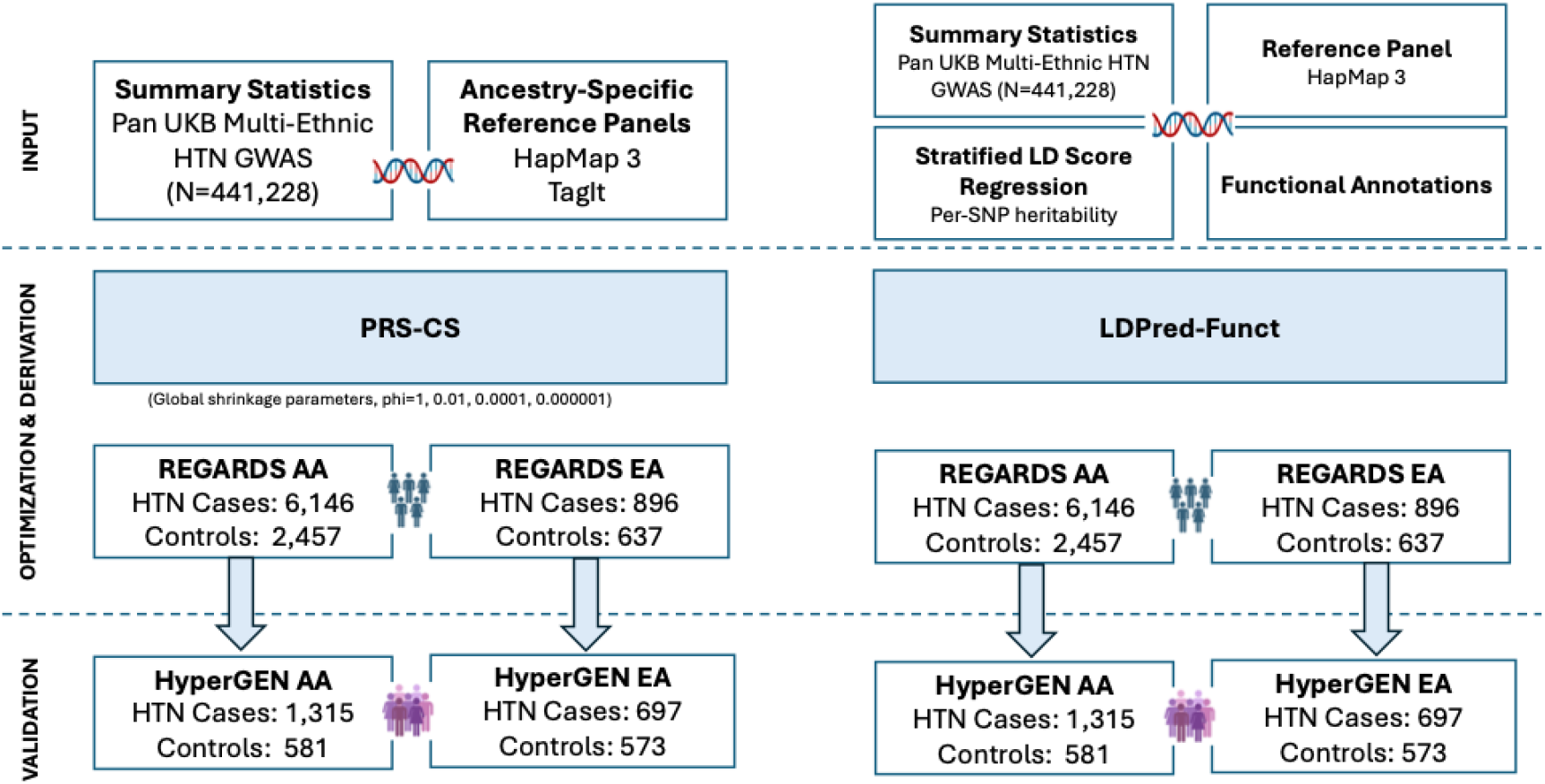
PRS Pipeline.

**Figure 2.**
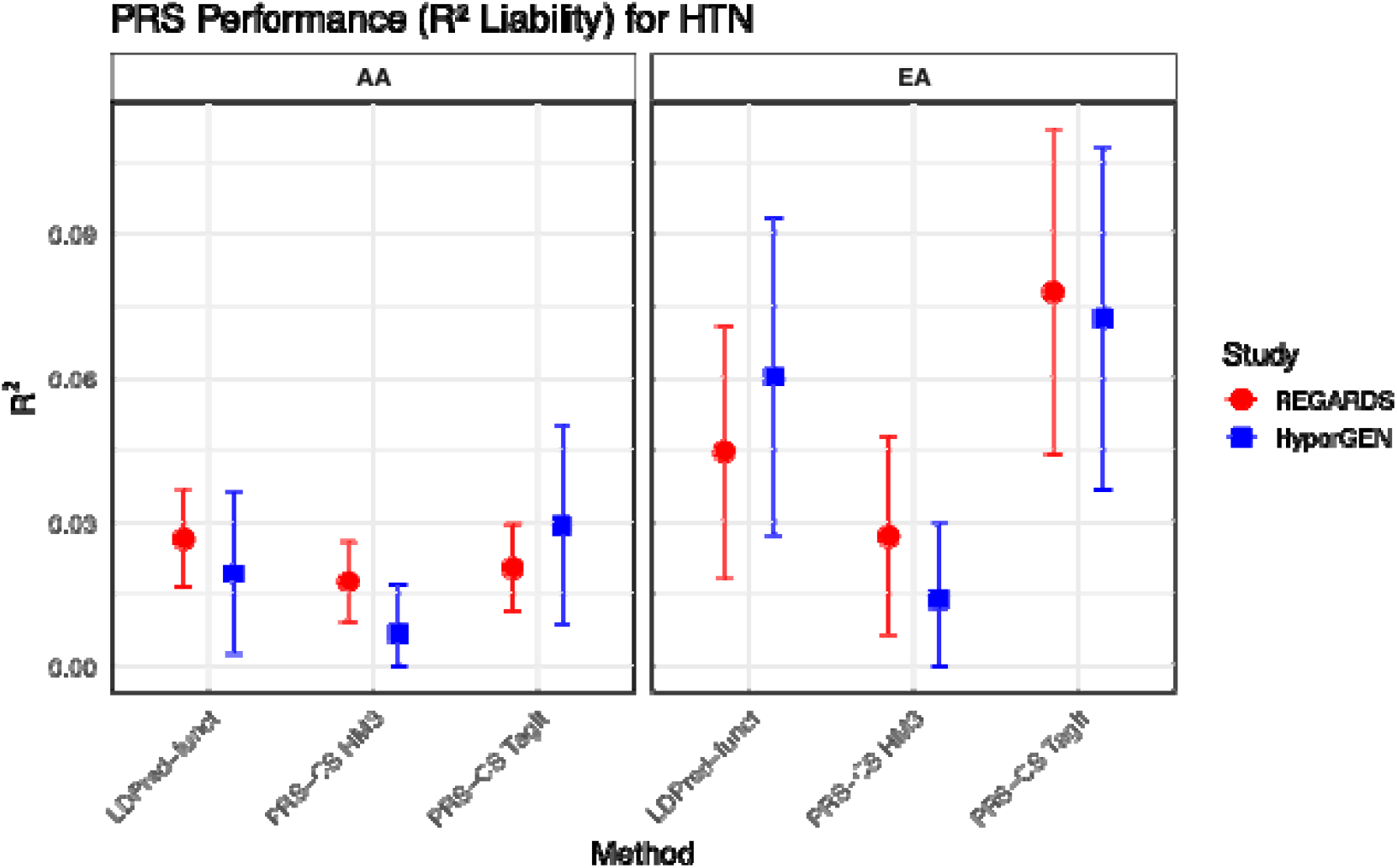
HTN R^2^-liability comparison by method per 1 SD of PRS.

In total, we generated 20 PRSs for HTN in the REGARDS data (10 for each ancestry group, where 4 (one for each ϕ) were output for PRS-CS-HM3, 4 (one for each ϕ) for PRS-CS-TagIt, and 2 (raw and p-value based) for LD-Pred-funct). The predictive performance of each PRS was assessed via a range of metrics. To measure the overall prediction accuracy, we calculated the proportion of variation in the trait explained by the PRS on the liability scale (R^2^ liability) after accounting for a basic set of covariates, including age, sex, and the top 10 ancestry principal components (PCs).(39, 40) Additionally, we calculated the area under the curve (AUC) for a covariates-only model (age, sex, and top 10 PCs), a PRS-only model, and a combined model (PRS plus covariates); and the odds ratio (OR) per standard deviation (OR/SD) change in the PRS, as well as the thresholded ORs (top 2%, 5%, or 10% of the PRS distribution relative to the remaining participants). Sensitivity, specificity, positive predictive value (PPV; the proportion of identified high-risk individuals who are true HTN case subjects), and negative predictive value (NPV; the proportion of unidentified individuals that are HTN control subjects) are also reported. Since PPV and NPV depend on the prevalence of the disease (we used 50% for both EAs and AAs), we report prevalence-adjusted PPV and NPV similarly to a previous report.(41) We tested all the same metrics for the 10 scores in each ancestry group in the HyperGEN cohort. All HyperGEN models were mixed models accounting for familial relatedness. For all PRS-CS models the best ϕ value score for REGARDS as determined by highest R^2^ are presented in the main tables and is the focus of the validation in HyperGEN.

## Data Availability

All data produced are available online on dbGAP.

https://dbgap.ncbi.nlm.nih.gov/beta/study/phs002719.v1.p1/#study

https://dbgap.ncbi.nlm.nih.gov/beta/study/phs001293.v3.p1/#study

---

**Supplemental Figure 1. AUC comparison for prediction of HTN**. A) AUC comparisons in AA participants from REGARDS optimization cohort. B) AUC comparisons in AA participants from HyperGEN validation cohort. C) AUC comparisons in EA participants from REGARD optimization cohort. D) AUC comparisons in EA participants from HyperGEN validation cohort

