## Supplementary figures and images for "Improving performance of polygenic risk scores for hypertension across two ancestry groups"

### Supplemental Figure 1

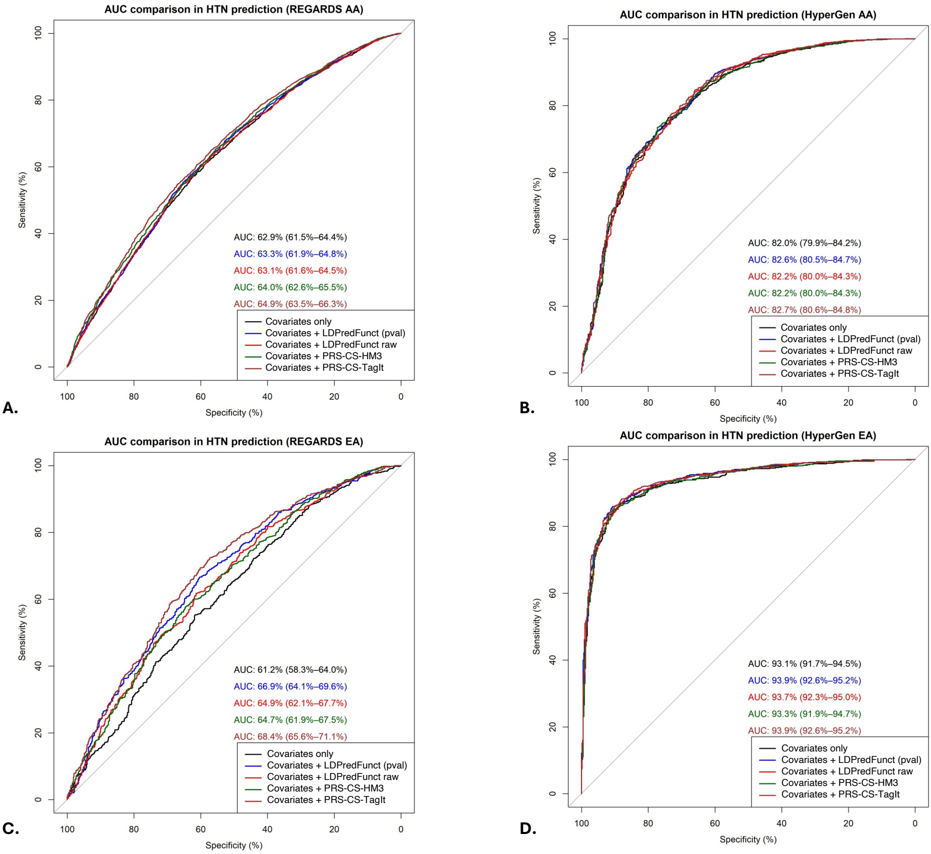
